# Cost-effectiveness of policy options for transformation of cytology-based nationwide cervical cancer screening programme in the Czech Republic: model-based economic evaluation

**DOI:** 10.64898/2026.02.12.26346126

**Authors:** Kateřina Hejcmanová, Ondřej Ngo, Renata Chloupková, Vladimír Dvořák, Markéta Trnková, Jaroslava Dušková, David Cibula, Ladislav Dušek, Karel Hejduk, Ondřej Májek

## Abstract

**Objectives:** Cervical cancer is a preventable disease, and a properly implemented screening programme can reduce its incidence and mortality and potentially save resources. This study aimed to evaluate the cost-effectiveness of options for potential transformation of the nationwide screening programme in the Czech Republic, especially considering recent changes in HPV DNA testing recommendations.

**Methods:** A microsimulation model was developed to assess the cost-effectiveness and health benefits of alternative screening strategies in the Czech Republic. The model simulated annual life cycles of women from age 15, comparing combinations of cytology and HPV testing. Input parameters used were obtained from national registries in the Czech Republic and from published literature. The analysis was conducted from the perspective of healthcare payers. Costs (2025 EUR) and LYs were discounted at a rate of 3% annually. Probabilistic sensitivity analysis was conducted.

**Results:** The CEA showed that, compared to the current setting (annual cytology with co-test at 35, 45, 55), only specific co-testing strategies lead to a decrease in incidence and mortality but differ in benefits and economic efficiency. The lowest ICER was reported for a strategy combining cytology at two-year intervals and co-testing at four-year intervals from ages 30 to 65. Sensitivity analysis showed that the current strategy has the highest probability of cost-effectiveness at €31,000 per LY gained. At higher values, this is replaced by a strategy with a 3-year interval co-test.

**Conclusions:** Based on the models presented, co-testing appears to be cost-effective. The actual willingness to pay threshold will facilitate selection of the most-appropriate strategy.

## Introduction

Cervical cancer is one of the most common cancers in women globally.^1^ When comparing the standardized mortality rate of the Czech Republic with other EU countries, there is still room for improvement. Based on available international data, the Czech Republic ranks among average EU countries in terms of standardized mortality from cervical cancer.^2^ Based on the national registries in the Czech Republic, approximately 750 women are diagnosed with this disease every year, and 300 die from the disease.^3^ However, cervical cancer is almost completely preventable, with the right combination of primary (vaccination) and secondary (screening) prevention.^1^ In case of screening, there are two most commonly used screening tests, which are cytology (Papanicolaou smear or liquid-based cytology) and human papillomavirus DNA test.^4^ In 2010 and 2015, the European Guidelines for Quality Assurance in Cervical Cancer Screening (and its supplements) were updated, and the main point was the recommendation to perform cervical cytology as a standard screening test from 20 to 65 years at 3- to 5-year intervals.^5,6^ Since then, a number of different studies have been published, and the HPV DNA test is recommended as the primary screening examination in many of them; some of them also recommend co-test (cytology and HPV DNA test) in defined time intervals.^7,8^ One of the documents that has recently determined the direction of cervical cancer screening in Europe is the 2022 *Council Recommendation on strengthening prevention through early detection*. This recommendation states that HPV testing should be used for women aged 30 to 65, at intervals of 5 years or longer.^9^ European guidelines on cervical cancer screening and diagnosis have also recently been published that support the introduction of HPV testing in primary screening.^10^

In 2008, an organised national cervical cancer screening programme was introduced in the Czech Republic. Standard screening examination was the Papanicolaou smear (Pap test) performed every year in women aged 15 and older. In 2021, the programme was extended to include HPV DNA testing in women aged 35 to 45, and from 2024 also in women aged 55 years.^11,12^ Introduction of HPV DNA testing in selected age groups should facilitate a comprehensive evaluation of the programme to analyse policy options for future programme strategy.

In every screening programme, it is necessary to monitor potential benefits and harms to the target population, but also the cost-effectiveness of the programme. This type of monitoring is particularly important when considering changes. Recently, studies have been published demonstrating the cost-effectiveness of incorporating HPV DNA testing into cervical cancer screening in specific health care systems.^13–15^ However, for many countries where cervicovaginal cytology has been the primary screening method for a long time, the transition to primary HPV DNA testing may be complicated, for example, because of the cost of testing equipment, setting up the testing laboratories, coordination, and transportation of the samples.^16^

In the context of the recent provisional changes in HPV DNA testing in the Czech Republic, this study aims to evaluate the cost-effectiveness of potential policy options for the cervical cancer screening programme in the Czech population and provide guidance and support materials for innovations of the screening strategy considering primary HPV DNA testing.

## Methods

### Model of the natural history of cervical cancer and screening strategies

A microsimulation model of the natural history of cervical cancer was created, the structure of this model was inspired by previously published models, especially the model published in Germany in 2006.^17,18^ The model consists of 11 natural history states: no cervical lesion or infection (well), high-risk HPV (hrHPV) infection, 3 precancerous states – cervical intraepithelial neoplasia (CIN) I-III (CIN III includes also adenocarcinoma in situ), 4 clinical states of the disease – stage I-IV, death from cervical cancer and death from other causes. The model assumes that cervical cancer cannot develop without a previous hrHPV infection. The modelled strategies are listed in Table 1. Another assumption is that in the case of co-testing, all women who undergo cytology also undergo HPV testing.

**Table 1.** Modelled strategies.

| Abbreviation | Description |
| --- | --- |
| Cyt15_1y | Annual cytology from 15 years |
| Cyt15_1y_CO35_45 | Annual cytology from 15 years, co-test* in 35 and 45 years |
| Cyt15_1y_CO35_45_55 | Annual cytology from 15 years, co-test* in 35, 45, and 55 years |
| Cyt15_1y_CO35_45_55_65 | Annual cytology from 15 years, co-test* in 35, 45, 55, and 65 years |
| Cyt15-29_1y_CO30-65_3y_Cyt3y | Annual cytology from 15 to 29 years, co-test* from 30 to 65 years every 3 years, followed by 3-year cytology from 66 years |
| Cyt15-29_1y_HP30-65_5y_Cyt5y | Annual cytology from 15 to 29 years, HPV DNA test from 30 to 65 years every 5 years, followed by 5-year cytology from 66 years |
| Cyt15_2y_CO30-65_4y | 2-year cytology from 15 years, co-test* from 30 to 65 years every 4 years |

\* co-test = cytology and HPV DNA test

Figure 1 describes a model of the natural history of cervical cancer with incorporated screening and detection of the clinical stages. Treatment for CIN II and III stages was also considered within the presented model. The model assumes that if these precancerous stages are detected, the woman will undergo conization. If a woman is diagnosed with CIN I (or HPV infection in strategies that use HPV DNA tests), she is monitored and, if there is no regression or persistence of the condition, she will also undergo conization when the condition progresses to CIN II or III.

**Figure 1.**
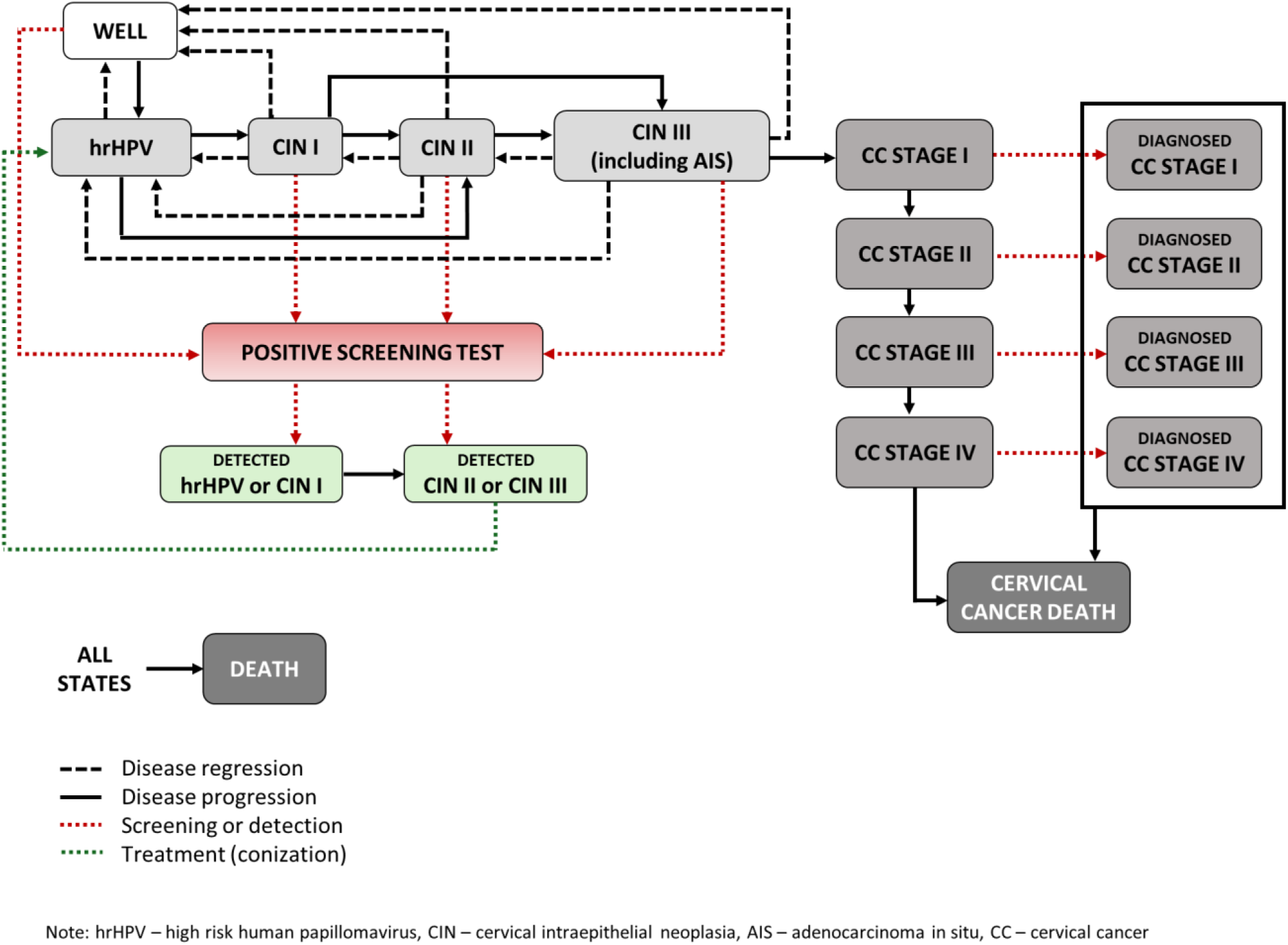
A model of the natural history of cervical cancer, including screening and detection of cervical cancer

### Model parameters, calibration, and simulation process

The advantage of simulation models is the ability to combine a variety of data sources. Some of the parameters were taken from the national registries and other available data sources in the Czech Republic. However, some of them are currently not available in the Czech context, and it was necessary to adopt them from international literature. For this reason, the overall model was calibrated to the Czech population after simulation. Table 2 demonstrates groups of parameters with appropriate source information. Parameters can be divided into two parts: input parameters, which are those that create the model and simulation. The second part are calibration and validation parameters, which are used for calibration to the Czech population and validation of the whole model.

**Table 2.** Simulation model parameters and their data sources.

| **Parameters** | **Data source** |
| --- | --- |
| ***Input parameters*** |  |
| Transition probabilities between states | Calibrated from published literature<sup>17,19–21</sup> |
| Five-year relative survival | Czech National Cancer Registry<sup>22</sup> |
| Probability of death from other causes | Complete Life Tables,<br>Czech Statistical Office<sup>23</sup> |
| Age-specific screening coverage | Czech National Registry of<br>Reimbursed Health Services<sup>24</sup><br>Dutch cervical cancer screening data<sup>25</sup> |
| Sensitivity of the screening examination | Calibrated from published literature<sup>17</sup> |
| ***Calibration and validation parameters*** |  |
| Age-specific incidence | Czech National Cancer Registry<sup>22</sup> |
| Age-specific mortality | Czech National Cancer Registry<sup>22</sup> |

As mentioned above, the assumed parameters were calibrated as part of the simulation process. For each parameter, the minimum and maximum value was set, based on the literature, and 1,000 combinations of these parameters were created by random selection from a uniform distribution for each parameter. A simulation was performed for each combination of parameters and the model output incidence was compared with the observed incidence in the Czech Republic (calibration parameter). The least square method was used for this comparison. Age-specific mortality was used as a validation parameter because it was not included in the model and was not used as a calibration target.

Every simulation was run on a sample of 100,000 women entering the model at the age of 15 years. Every woman started in a state of full health. One cycle describes one year of a woman’s life, and the model has 85 cycles (it models women aged 15 to 100 years or death). Each woman could move between states according to the arrows shown in Figure 1 or stay in the same state for several cycles. The model outcome is age-specific incidence and mortality, number of person-years of life, and number of performed screening examinations (cytology and HPV test). If the woman is in any state other than death, 1 person-year of life is considered. Health benefits are assessed as changes in age-specific incidence and mortality, but also as the number of person-years gained.

### Lifetime cost of cervical cancer care and cost of screening

In order to compare the costs incurred, it was necessary to estimate the cost of care for women with cervical cancer. We utilised official Czech national healthcare data from the National Health Information System allowing for secondary data use.^26,27^ Women with cervical cancer were identified in the Czech National Cancer Registry (CNCR)^22^. Subsequently, women who were prevalent in the 2018-2022 period were followed for all covered health services. All those that are in the Czech National Registry of Reimbursed Health Services (NRRHS), and this part of the analysis deals with actual reimbursed expenses.^24^ Data from the start of the follow-up (1/1/2018) to the death or the end of the year 2022 were considered. The costs were stratified by age of the women, diagnosed stage, and clinical phase (initial, continuous, and terminal). The initial phase represented the first 12 months from the date of diagnosis. The terminal phase represented the last 12 months of life. The continuous phase represented the entire period between the initial and terminal phases. If a person did not live with the diagnosis for more than 12 months, the entire period was considered the terminal phase. Subsequently, the initial phase was considered.^28^

A control group of women without the observed disease was identified by random selection and subsequent matching based on the year of birth. Healthy women were assigned a pseudo date of diagnosis, according to the matching, to identify the start of reimbursed health services tracking. The average annual costs of women without the disease were subtracted from the average annual costs of women with cervical cancer. The CNCR was used to determine the lifetime cost of care for patients diagnosed with cervical cancer, specifically the overall survival of women with cervical cancer by stage.

Unlike the previous section, which is based on actual costs, screening costs are entered into the analysis as values reported in the Decree of the Ministry of Health. Decree No. 320/2023 Coll. was used for the analysis, which is the current version for the year 2024.^29^ The costs are further specified in the Decree No. 319/2023 Coll., which was also used.^30^ All costs included in the model are listed in the Supplementary material (Supplementary Tables 1a-1b).

### Model outcomes and cost-effectiveness analysis

For each modelled strategy, incidence, mortality, number of conizations provided, total costs (screening costs, including conization, and care for women with cancer), and person-years were calculated. When comparing screening strategies, only those that are currently more advanced than the strategy currently used in the Czech Republic (i.e., annual cytology from 15years, co-testing at ages 35, 45, and 55) were considered. An alternative calculation related to the basic strategy is provided in the Supplementary material. This alternative calculation considers as a baseline comparator a strategy with annual cytology without concurrent primary use of HPV testing.

The outcome of the model for the compared strategies was a change in incidence and mortality, a change in conizations provided, incremental costs, and person years gained. Based on those outcomes, the incremental cost-effectiveness ratios (ICER) were calculated to finally compare alternative screening strategies.

The entire model was created from the perspective of a healthcare payer, i.e., health insurance companies. All costs are expressed in 2025 Euros, costs and LYs were discounted at a rate of 3% per year.

### Sensitivity analysis

Of all the parameters considered for inclusion in the model, a basic set of parameters with the highest degree of uncertainty was selected. Since the parameters for the natural transition between states were taken from the published literature, in many cases based on clinical studies, and were further calibrated to the Czech population, they were not evaluated in the sensitivity analysis. As part of the sensitivity analysis, cost values were varied based on expert estimates, and threshold values of ± 20% were set. Furthermore, the values of coverage by screening test and annual probability of death from cervical cancer were varied – for all these parameters, a 95% confidence interval (CI) was chosen based on observed data in the Czech Republic. A probability sensitivity analysis (PSA) was performed. A set of 1,000 randomly selected sets of data parameters was created and simulated in all considered strategies. A uniform distribution was assumed for all monitored parameters. Incremental costs and LYs gained were calculated for each set. For each simulation, the net monetary benefit (NMB) was then calculated, based on the range of potential willingness to pay (WTP) thresholds. Based on the results obtained, a cost-effectiveness acceptability curve (CEAC) was plotted.

### Implementation of the model

The model was created in R Statistical Software version 4.0.5 and later updated in version 4.2.3, specifically in the development environment R Studio.^31^ The model was created using a tutorial published in 2018.^32^ The model can be obtained on the Zenodo web platform and used for further purposes and modelling.^33^

## Results

### Input parameters and baseline calibrated model

All the transition probabilities between states were derived from published literature and calibrated to the Czech population. The set of parameters that was evaluated as the best is listed below in the Supplementary material (Supplementary Tables 2a-2c). The remaining parameters calculated for model inputs and listed in the Supplementary material were obtained from individual national registries. Data on cervical cancer screening coverage of the target population in the Czech Republic are available for 1-year, 2-year, 3-year, and 5-year intervals (Supplementary Table 3). The coverage of women by screening examinations was therefore used based on NRRHS data, except for the model that works with a solo HPV test between the ages of 30 and 65. In this case, a completely different screening strategy is required, and the current 5-year coverage figures cannot be applied. For this reason, Dutch cervical cancer screening data was used, where the transition to primary HPV testing has already taken place. Overall participation there is around 70%, but age-specific data is entered into the model.^25^ Further data show the probability of women dying from any cause by age (Supplementary Table 4). The last table of input parameters shows five-year relative survival for cervical cancer and derived annual probability of death by cervical cancer stage (Supplementary Table 5).

Using the least square method, the calibration process selected a set of input transition probabilities (parameters) that best fit to the population of women in the Czech Republic. Validation of the outcome of this model was performed on age-specific mortality. Observed curves and best-fit model incidence and mortality are presented in Supplementary materials (Supplementary Figures 1a-1b).

### Health benefits and cost-effectiveness of screening strategies

Table 3 shows all outcomes of the modelled strategies and a comparison with the strategy currently used in the Czech Republic (strategies compared to the model *Cyt15_1y_CO35_45_55*). An alternative comparison with the annual cytology strategy is provided in the Supplementary material (Supplementary Table 6). The models were ranked in ascending order according to the costs incurred, and only one of the strategies (*Cyt15-29_1y_HPV30-65_5y_Cyt5y*) was evaluated as dominated. Figure 2 shows the cost-effectiveness plane of screening strategies.

**Figure 2.**
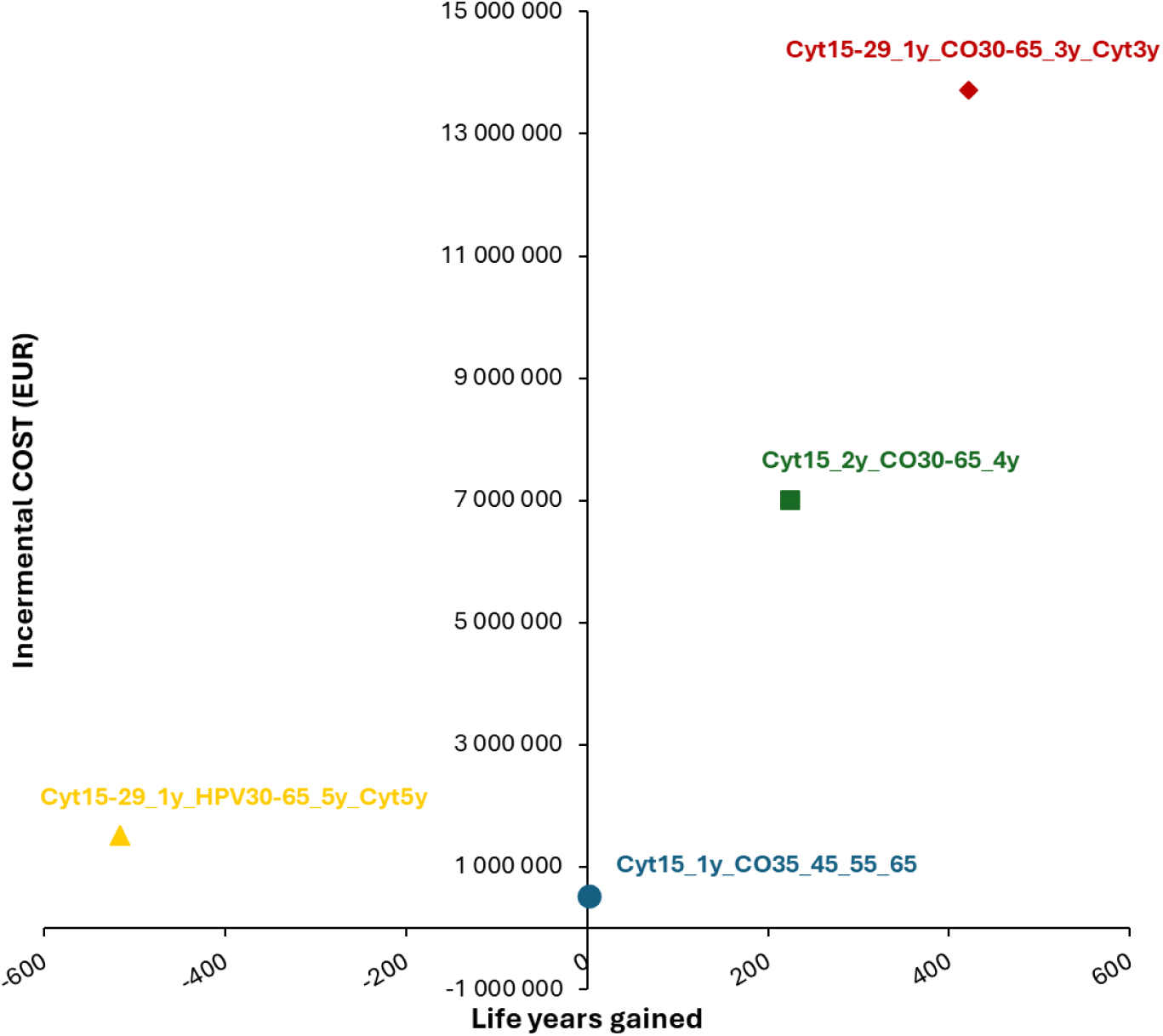
Cost-effectiveness plane of simulated screening strategies

**Table 3.**
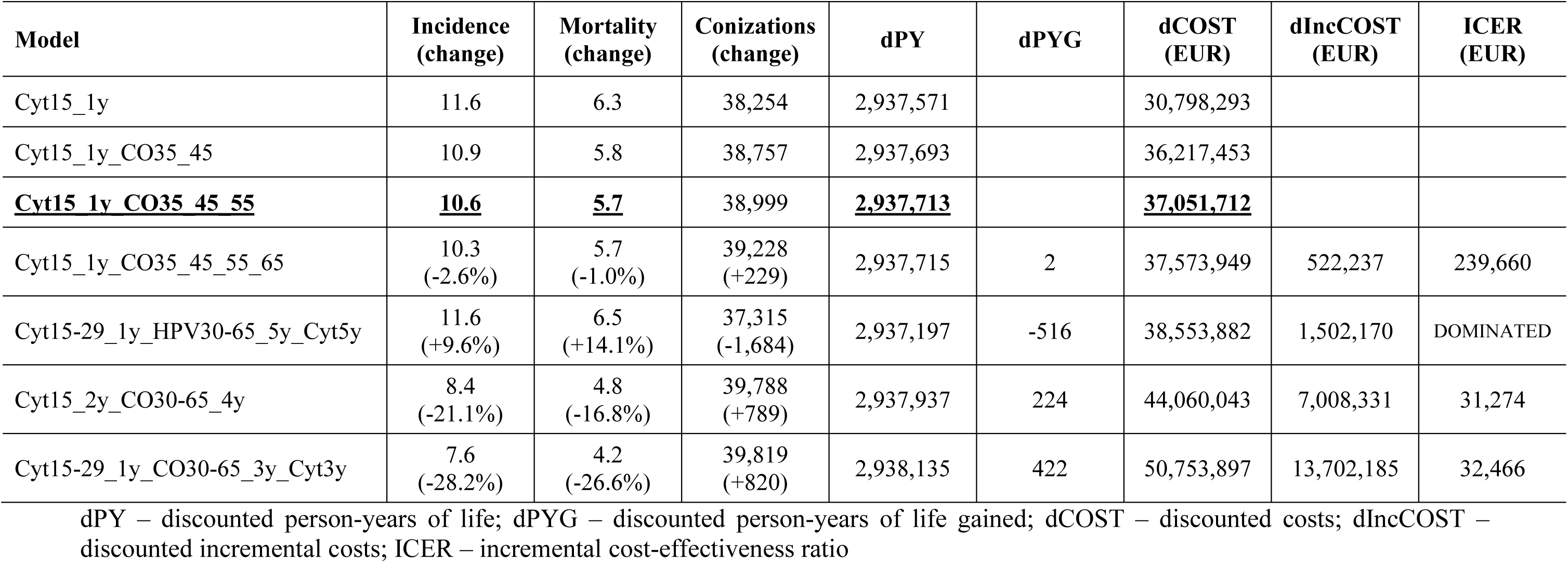
Model benefits and costs of strategies.

A decrease in incidence and mortality can be observed in all of the strategies compared (except the dominated). With increasing costs, we observe an increasing number of life years gained. The lowest ICER is observed in strategy *Cyt15_2y_CO30-65_4y*, which is also associated with a increase in the number of conizations performed. The decrease in incidence and mortality in this case is 21.1% and 16.8%, respectively. Strategy *Cyt15-29_1y_CO30-65_3y_Cyt3y* shows a similar ICER (31,274 vs. 32,466 EUR) and a similar number of conizations performed (39,788 vs. 39,819). Differences can be seen in affordability (increase of the incurred cost) and in the decline in incidence and mortality, with strategy *Cyt15-29_1y_CO30-65_3y_Cyt3y* showing a greater decline. In contrast to the above results, strategy *Cyt15_1y_CO35_45_55_65* shows a high ICER and only a slight decline in incidence and mortality. The ineffectiveness of this strategy is likely due to the relatively low screening coverage among women aged 65, to whom an HPV test is added to routine cytology in the strategy under review (see Supplementary Table 2).

### Sensitivity analyses

As part of the sensitivity analysis, the parameters of costs, screening coverage of women, and the probability of death from cervical cancer were varied. PSA was performed with 1,000 sets of randomly selected data parameters. Given that there is currently no threshold for WTP in screening interventions in the Czech Republic, results were monitored for thresholds ranging from 0 to 60,000 EUR per LY gained. Figure 3 shows the CEAC based on the WTP values. Up to a value of 31,000 EUR per LY gained WTP, current strategy *Cyt15_1y_CO35_45_55* has the highest probability of cost-effectiveness, but this probability decreases as the WTP value increases. From a value of 32,000 per LY gained, strategy *Cyt15-29_1y_CO30-65_3y_Cyt3y* takes over and achieves the highest probability. Subsequently, this strategy prevails with the highest probability, reaching 100% at higher values of WTP.

**Figure 3.**
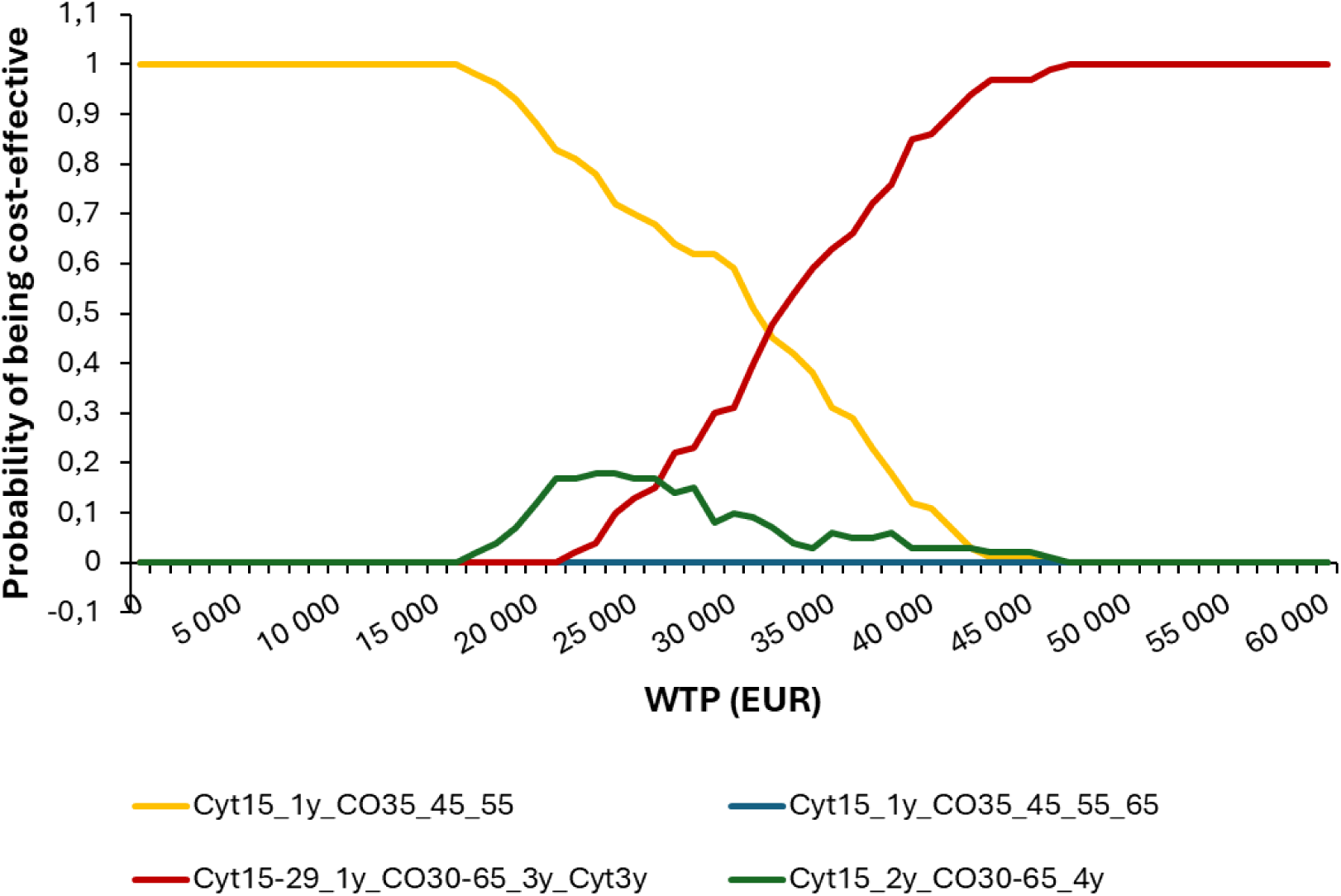
Cost-effectiveness acceptability curve

## Discussion

Cervical cancer screening is a long-established organised programme in the Czech Republic. This publication supports an organised change in the use of primary screening, which is currently being addressed by several European Union countries, and which was supported by an EU Council Recommendation. The results of the above models indicate a potential reduction in incidence and mortality if HPV testing is introduced into the routine screening process in the Czech Republic. Strategy *Cyt15_2y_CO30-65_4y* appears to be the most cost-effective, as it has the lowest ICER and also significantly reduces incidence and mortality. It is important to note, that unlike strategy *Cyt15-29_1y_CO30-65_3y_Cyt3y*, for which a very similar ICER was estimated (31,274 vs. 32,466 EUR), this strategy is less costly (44 vs 51 mil EUR). Conversely, strategy *Cyt15_1y_CO35_45_55_65* shows a high ICER with only a small impact on health benefits, which can be explained, among other things, by the low screening coverage in older age groups, where the HPV test is included in this strategy.

The results of the sensitivity analysis show that the probability of individual strategies being cost-effective depends heavily on the WTP threshold, which has not yet been established in the Czech Republic for screening interventions. At lower WTP values (specially up to 31,000 EUR per LY gained), strategy *Cyt15_1y_CO35_45_55* (current strategy) achieves the highest probability of cost-effectiveness. While for values above 31,000 EUR per LY gained, strategy *Cyt15-29_1y_CO30-65_3y_Cyt3y* begins to prevail, reaching a 100% probability at higher values. These results show that the parameters varied in these simulations can have a significant impact on the cost-effectiveness of individual strategies. While strategy *Cyt15_2y_CO30-65_4y* appeared to be the best (lowest ICER) in the point estimate, strategy *Cyt15-29_1y_CO30-65_3y_Cyt3y* was evaluated as the best in the sensitivity analysis from the threshold of 31,000 EUR.

Another important point of discussion is the determination of the WTP threshold. In the Czech Republic, this threshold has not yet been set for interventions in the form of screening programmes. However, several outputs show a value for pharmacoeconomic purposes of 48,000 EUR per QALY. In this context, of the model strategies described above, only *Cyt15_2y_CO30-65_4y* and *Cyt15-29_1y_CO30-65_3y_Cyt3y* can be considered cost-effective, while in the case of strategy *Cyt15_1y_CO35_45_55_65*, the ICER values lies above the potential WTP threshold in the Czech Republic.

Back to the results of the model, comparable findings were achieved in other European studies. In their study examining the optimal HPV screening protocol in Eastern Europe, Jansen et al. report that, based on the MISCAN-Cervix microsimulation model, cytology at ages 25 and 28, followed by a 5-year HPV test from ages 30 to 65, was evaluated as the most appropriate strategy for Slovenia.^34^ Similar results can also be seen in publications from Western Europe, such as Sweden and the Netherlands, where HPV testing has already been incorporated into routine screening programmes.^14,35^ The Swedish study also focused on the unvaccinated female population and, using mathematical modelling, presents as the most appropriate screening strategy one based on primary HPV testing at 5-year intervals for women aged 23 to 50, with the interval subsequently extending to 10 years from the age of 50.^14^ Similarly, a study from Germany also reports that a screening programme based on HPV testing is cost-effective in all variants of the sensitivity analysis considered. ^35^

The analysis described has a number of strengths. One of the most significant is the realistic estimate of the treatment costs for cervical cancer, which are based on individual data reported by healthcare payers. The model also considers long-term trends in survival rates for the monitored disease and works with highly accurate estimates of screening coverage among the female population, parameters that can significantly influence the results of the modelled strategies. Another advantage of this study is its local and contextual adaptability, as well as its generalizability. The study deals with the current situation in the Czech Republic, where cytological smear analysis is used as the primary screening strategy in the long term. The modelled strategies are thus adapted to this situation. On the other hand, the above models describe various stages of gradual transition from primary cytological screening to primary HPV screening (or co-testing), which is an issue currently being addressed by a number of European countries, and this study may thus be a useful tool for their decision-making.

This analysis has important limitations that should be noted. Although the models describe in detail the impact on the population of women, they do not consider the vaccination against HPV infection, which is currently on the rise, not only in the Czech Republic. However, it is important to take into account that vaccination of women covered by public health insurance started in the Czech Republic in 2009, and the first systematically vaccinated women are now aged 30. Future research should therefore focus, among other things, on an organised transition to screening strategies that consider the increasing proportion of women vaccinated against HPV. At this point, the presented output focuses only on the screening effect. It can be assumed that vaccination will not significantly affect the population burden of cervical cancer in the short-term future time. Secondly, the availability of data is also a limitation, especially in the case of transition probabilities between states of the natural history of the disease, which had to be taken from the published literature. Although these parameters were calibrated and validated on the Czech population of women (using epidemiology data), it is necessary to acknowledge some imprecision in these outputs. Another, equally important limitation is that the model assumes that in the case of a co-test, the HPV DNA test is performed on all women who have cytology performed. Available data from health insurance companies in the Czech Republic show that not all gynaecologists follow this practice, and only about 70% of women who should be co-tested actually have both tests. However, in the model, we assume that the HPV test is performed on all women in the case of co-testing.

## Conclusions

Based on the model estimates, the gradual introduction of HPV testing as part of primary screening for cervical cancer appears to be cost-effective, whether in the form of HPV testing alone in the 30-65 age group or in the form of co-testing. All modelled strategies point to significant benefits for the monitored population of women, including a reduction in incidence and mortality. On the other hand, there is an increase in the number of conizations performed. These results thus support the EU Council Recommendation, which supports the use of HPV primary testing in women aged 30 to 65 at 5-year intervals.

The results of this study provide information for supporting the decision on appropriate future cervical screening strategy for the Czech Republic and other countries designing the strategy for transition to HPV-based organised cervical cancer screening.

## Data Availability

Computational script is available at Zenodo.
Agregated data are included in the supplementary material.

https://doi.org/10.5281/zenodo.18611243

## Acknowledgements

The authors thank Ing. Eva Nevrtalová, Ph.D. for her valuable consultations and Mgr. Kateřina Opatřilová for assistance with data analysis. The work was supported from Programme JAC - project SALVAGE (CZ.02.01.01/00/22_008/0004644) financed by the Ministry of Education, Youth and Sports – Co-funded by the European Union. The authors also thank the RECETOX Research Infrastructure (No LM2023069) financed by the Ministry of Education, Youth and Sports for supportive background.

## Supplementary material

The Supplementary Tables 1a-1b show individual costs that are included in the cost-effectiveness analysis. Table 1a contains the average observed lifetime costs of care for women diagnosed with cancer according to stadium. Table 1b contains values of the costs of individual screening examinations.

**Supplementary Table 1a.** Observed costs lifetime of care for women diagnosed with cervical cancer.

| **Cervical cancer stage** | **Cost (EUR)** |
| --- | --- |
| Stage I | 31,115 |
| Stage II | 37,022 |
| Stage III | 37,816 |
| Stage IV | 21,109 |

**Supplementary Table 1b.** Screening costs.

| **Examination** | **Cost (EUR)** |
| --- | --- |
| Cytology | 14.5 |
| HPV DNA test | 55.8 |
| Conization | 77.7 |

The Supplementary Tables 2a-2c show calibrated values (2a: annual transition probabilities, 2b: annual probabilities of detection cervical cancer by symptoms, 2c: sensitivity of screening cytology) selected using the method of least squares with the references.

Annual probabilities represent values that have been calibrated for the Czech population. References represent input sources of values and their potential ranges, which were subsequently calibrated.

**Supplementary Table 2a.**
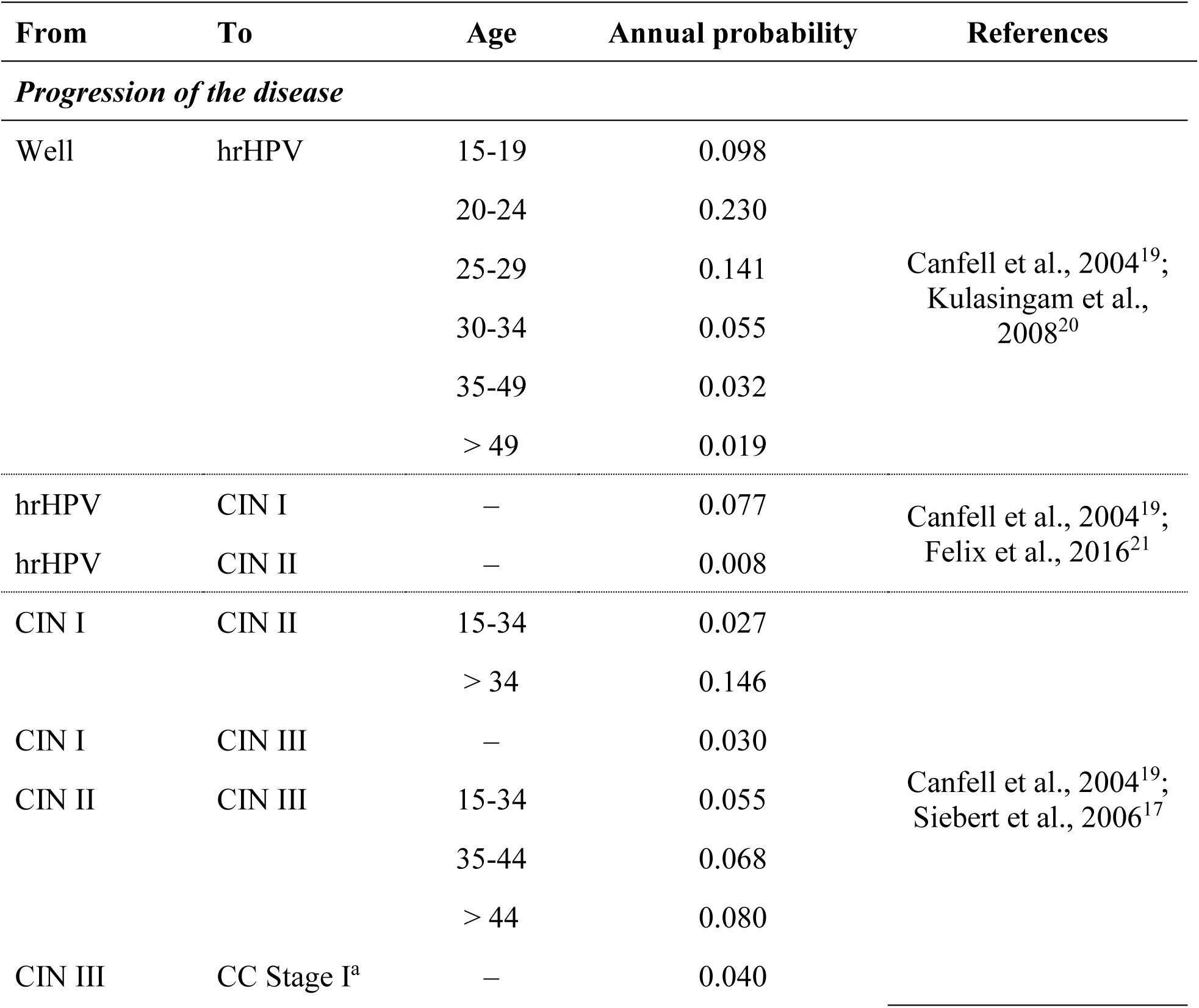
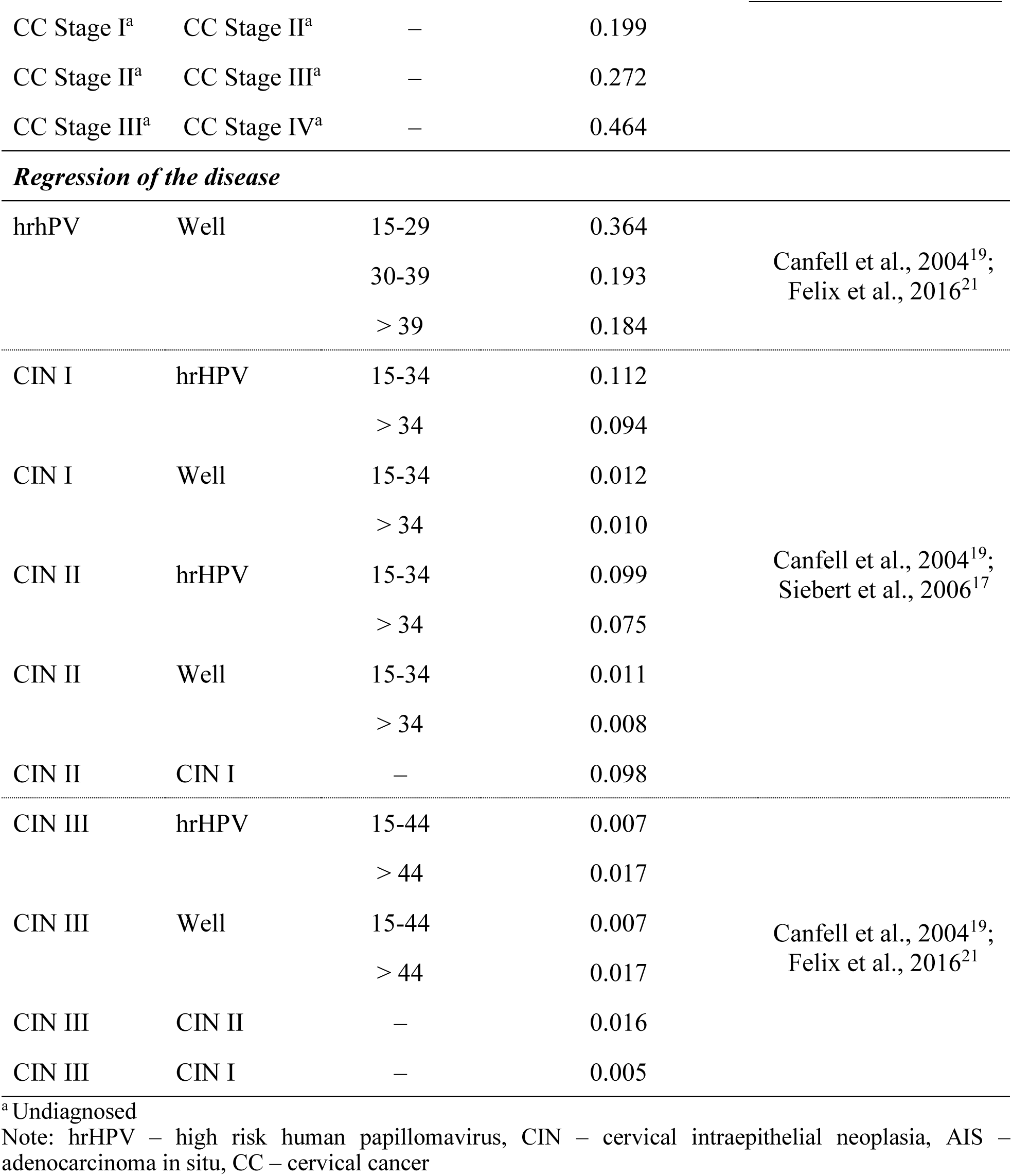
Calibrated transition probabilities used in microsimulation model – rounded up.

**Supplementary Table 2b.**
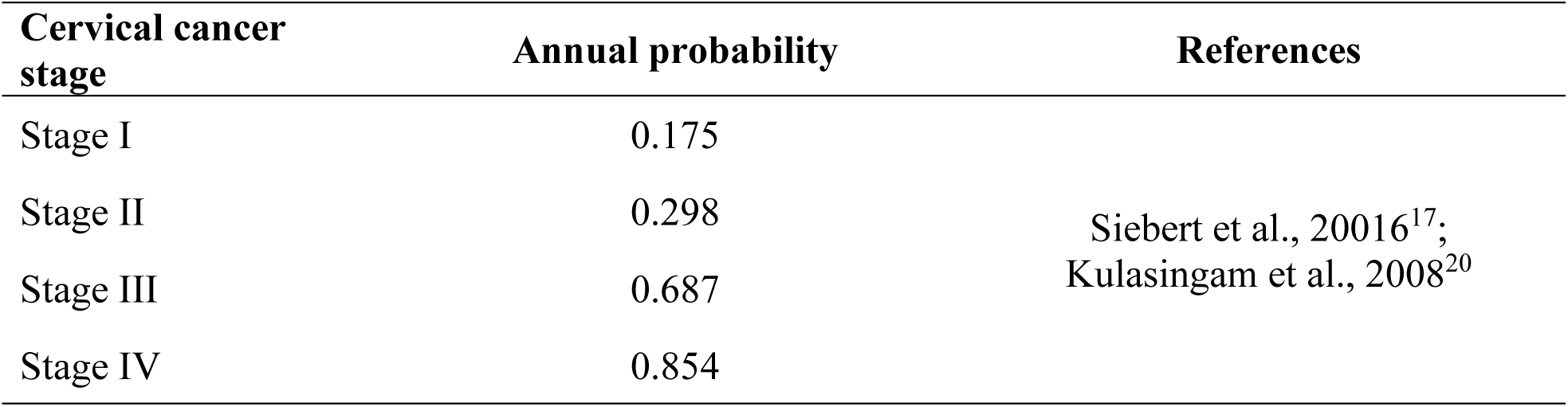
Calibrated probabilities of detection of cervical cancer by symptoms.

**Supplementary Table 2c.** Calibrated values of cervical cancer cytology sensitivity.

| **Stage** | **Sensitivity** | **References** |
| --- | --- | --- |
| CIN I | 0.467 | Siebert et al., 2006<sup>17</sup> |
| CIN II, CIN III | 0.744 |  |

Note: CIN – cervical intraepithelial neoplasia

The Supplementary Table 3 shows the cervical cancer screening coverage at one-year (1y), three-year (3y) and five-year (5y) interval by age (based on the data from 2022, calculated in 2023 from the National Registry of Reimbursed Health Services).

**Supplementary Table 3.**
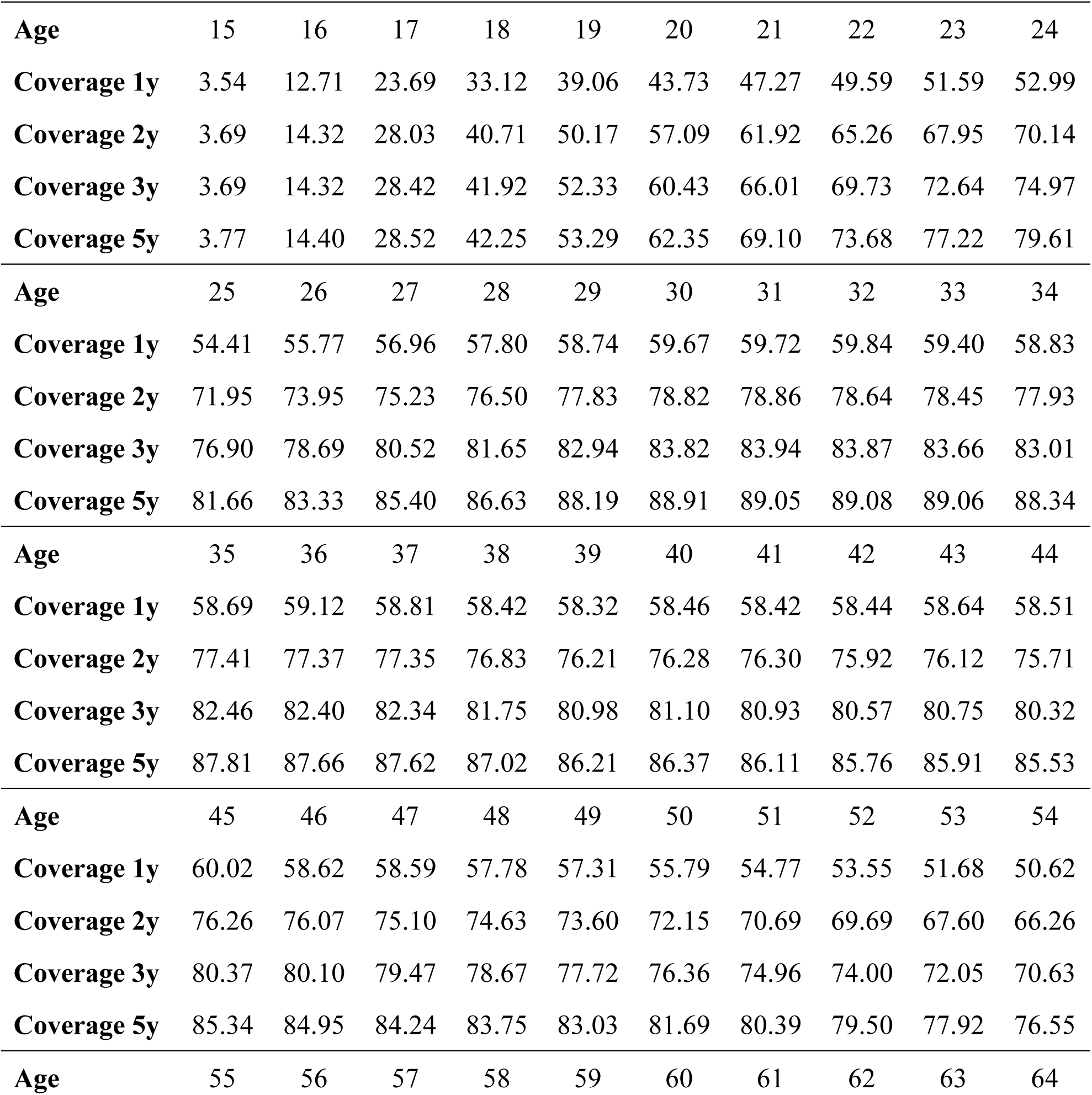
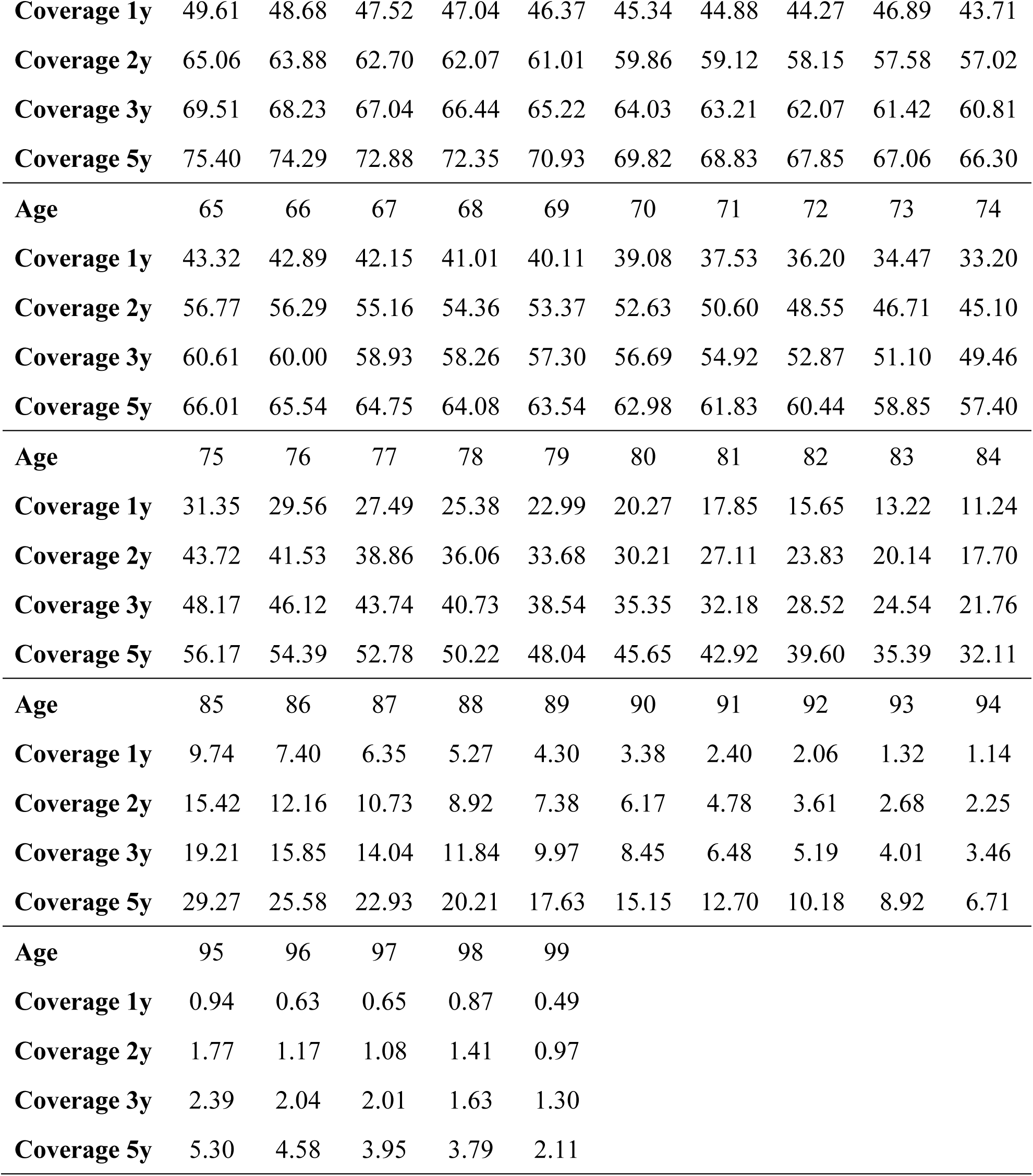
Cervical cancer screening coverage at one-year (1y), three-year (3y) and five-year (5y) interval by age (%) – rounded up.

The Supplementary Table 4 shows the probability of women dying from any cause by age (based on the data from 2022, published in 2023 by the Czech Statistical Office).

**Supplementary Table 4.**
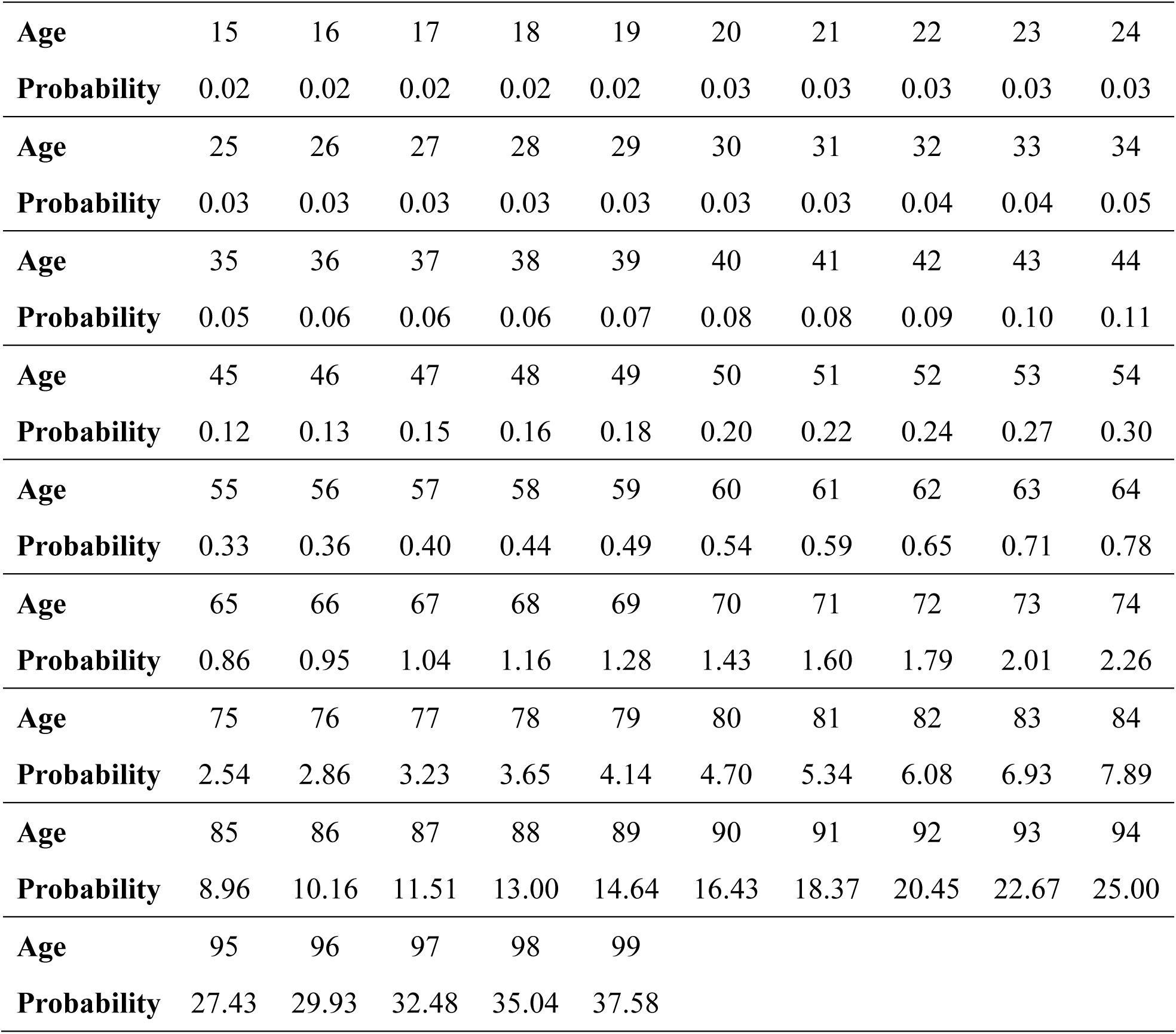
Probability of women dying from any cause by age (%) – rounded up.

The Supplementary Table 5 shows five-year relative survival for cervical cancer and derived annual probability of death (assuming an exponential distribution) by cervical cancer stage (based on the data from 2020-2022, calculated in 2023 from the Czech National Cancer Registry).

**Supplementary Table 5.** Five-year relative survival for cervical cancer and derived annual probability of death by cervical cancer stage – rounded up.

| **Cervical cancer stage** | **Five-year relative survival** | **Annual probability of death** |
| --- | --- | --- |
| Stage I | 0.918 | 0.017 |
| Stage II | 0.714 | 0.065 |
| Stage III | 0.546 | 0.114 |
| Stage IV | 0.183 | 0.288 |

The Supplementary Table 6 shows an alternative comparison of the cost-effectiveness of screening strategies compared to the basic strategy, i.e., annual cytology from 15 years of age.

**Supplementary Table 6.** Model benefits and costs of strategies (alternative comparison with basic strategy)

| Model | Incidence<br>(change) | Mortality<br>(change) | Conizations<br>(change) | dPY | dPYG | dCOST<br>(EUR) | dIncCOST<br>(EUR) | ICER<br>(EUR) |
| --- | --- | --- | --- | --- | --- | --- | --- | --- |
| Cyt15_1y | <u>11.6</u> | <u>6.3</u> | <u>38,254</u> | <u>2,937,571</u> |  | <u>30,798,293</u> |  |  |
| Cyt15_1y_CO35_45 | 10.9<br>(-6.2%) | 5.8<br>(-7.5%) | 38,757<br>(+503) | 2,937,693 | 123 | 36,217,453 | 5,419,160 | 44,185 |
| **<u>Cyt15 1y CO35 45 55</u>** | 10.6<br>(-8.8%) | 5.7<br>(-8.8%) | 38,999<br>(+745) | 2,937,713 | 142 | 37,051,712 | 6,253,418 | 43,974 |
| Cyt15_1y_CO35_45_55_65 | 10.3<br>(-11.2 %) | 5.7<br>(-9.8%) | 39,228<br>(+974) | 2,937,715 | 144 | 37,573,949 | 6,775,656 | 46,927 |
| Cyt15-29_1y_HP30-65_5y_Cyt5y | 11.6<br>(-0.1%) | 6.5<br>(+4.0%) | 37,315<br>(-939) | 2,937,197 | -374 | 38,553,882 | 7,755,589 | DOMINATED |
| Cyt15_2y_CO30-65_4y | 8.4<br>(-28.1%) | 4.8<br>(-24.1%) | 39,788<br>(+1,534) | 2,937,937 | 366 | 44,060,043 | 13,261,749 | 36,204 |
| Cyt15-29_1y_CO30-65_3y_Cyt3y | 7.6<br>(-34.5%) | 4.2<br>(-33.1%) | 39,819<br>(+1,565) | 2,938,135 | 564 | 50,753,897 | 19,955,604 | 35,366 |

dPY – discounted person-years of life; dPYG – discounted person-years of life gained; dCOST – discounted costs; dIncCOST – discounted incremental costs; ICER – incremental cost effectiveness ratio

The Supplementary Figure 1a-1b shows age-specific incidence and mortality. Blue line shows observed parameters in recent period. Red line shows simulated parameters. The simulated incidence and mortality are outcomes of the model, which was evaluated as the best using the least square method. Incidence was chosen as calibration parameter, mortality as validation parameter.

**Supplementary Figures 1a-1b.**
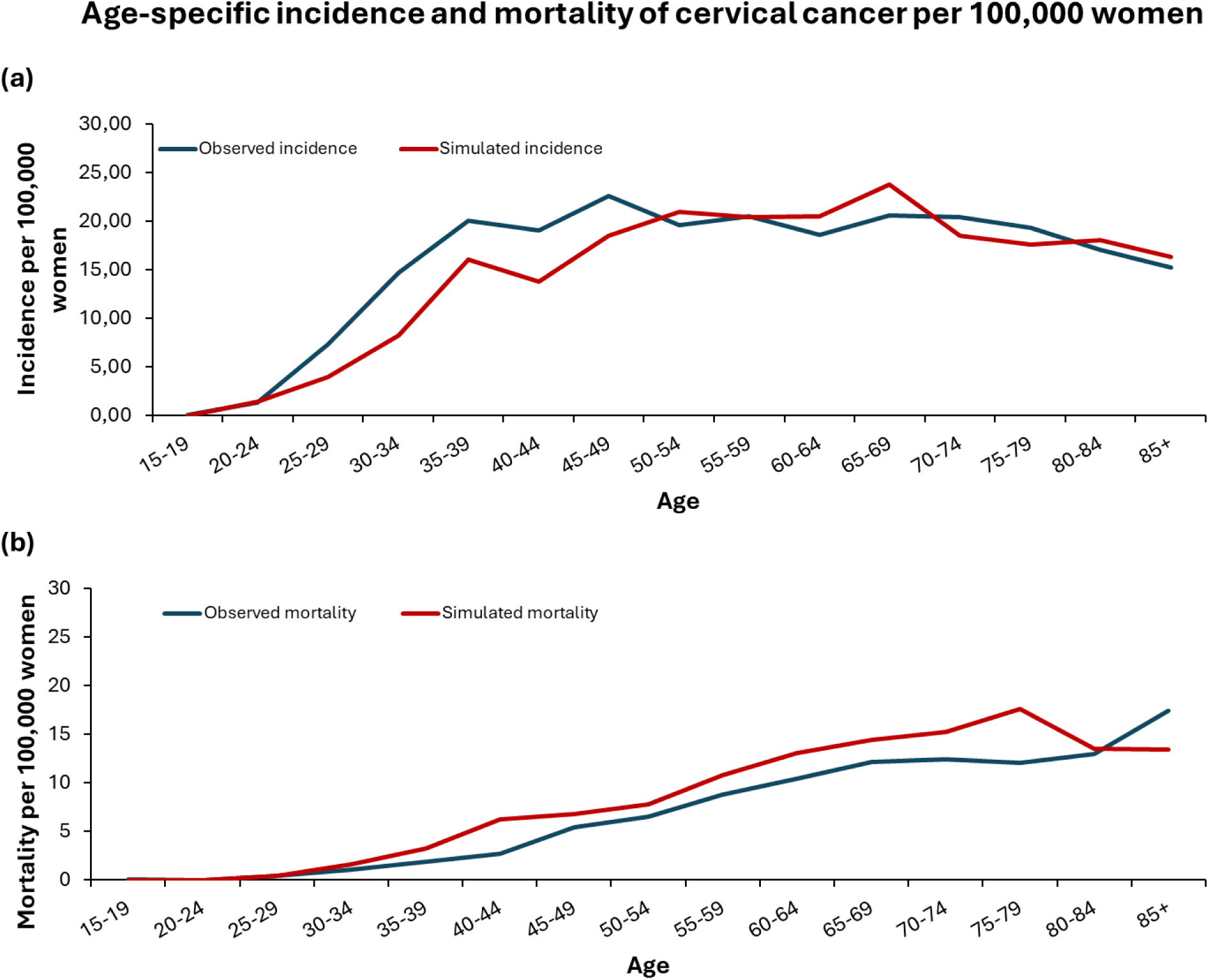
Age-specific incidence (a) and mortality (b) of cervical cancer per 100,000 women – observed and simulated values

